# A COVID-19 Risk Assessment for the US Labor Force

**DOI:** 10.1101/2020.04.13.20063776

**Authors:** Samantha Maher, Alexandra E Hill, Peter Britton, Eli P. Fenichel, Peter Daszak, Carlos Zambrana-Torrelio, Jude Bayham

## Abstract

The consequences of COVID-19 infection varies substantially based on individual social risk factors and predisposing health conditions. Understanding this variability may be critical for targeting COVID-19 control measures, resources and policies, including efforts to return people back to the workplace. We compiled individual level data from the National Health Information Survey and Quarterly Census of Earnings and Wages to estimate the number of at-risk workers for each US county and industry, accounting for both social and health risks. Nearly 80% of all workers have at least one health risk and 11% are over 60 with an additional health risk. We document important variation in the at-risk population across states, counties, and industries that could provide a strategic underpinning to a staged return to work.

**One Sentence Summary:** There is important variability in the proportion of the US workforce at risk for COVID-19 complications across regions, counties, and industries that should be considered when targeting control and relief policies, and a staged return to work.

## Main Text

The severity of the current COVID-19 epidemic in the US will depend on biological and epidemiological factors related to the SARS-CoV-2 virus, behavioral response of human populations including adherence to physical (social) distancing, the public health response, and policy makers’ ability to set and enforce control measures. COVID-19 is a novel disease in the US and is known to be transmitted by symptomatic and asymptomatic carriers *(1)*. In the US, COVID-19 has already caused significant public health burden in urban centers, and two-thirds of rural counties are now reporting cases *(2, 3)*. The consequences of infection on individuals are highly variable *(4)*, with most cases being mild, and approximately 20% of reported cases becoming severe *(5, 6)*. The severity of disease outcome is not random, and a number of predisposing conditions and risk factors are disproportionately linked to hospitalization and death [Table 1]. Currently, most suspected cases in the US are only admitted to hospital when severe (e.g. due to difficulty in breathing), because of anticipated overburdening of hospital capacity, medical staff, PPE, ventilators and ICU beds. Here, we use administrative data from the US National Health Interview Survey (NHIS)*(7)* and the US Quarterly Census of Earnings and Wages (QCEW) *(8)* to identify which groups of the labor force are at heightened risk of COVID-19 hospitalization and death. Our results are visualized for each county by industry and risk factor, and can be viewed using an interactive map at https://covid.yale.edu/resources/complications/. This information is important because high rates of complications in critical industries can disrupt the provision of goods and services.

**Table 1:** Breakdown of Risk Variables

| Health Risk Variables |  |  |
| --- | --- | --- |
| Variable Name | Description | Literature |
| Any risk | People with a health risk factor of all ages | Aggregate variable |
| Asthma | Currently has asthma | (8) |
| BMI > 25.5 | Has a BMI greater than 25.5 | (19, 20) |
| Cancer | Has ever had cancer | (21) |
| Diabetes | Ever told had diabetes | (22-24) |
| Heart Condition | Ever told had heart condition/disease | (21, 22, 25) |
| Hepatitis | Ever had hepatitis | (26) |
| Hypertension | Had hypertension, past 12 months | (21, 27) |
| Kidney Disease | Told had weak/failing kidneys, past 12 months | (25) |
| Liver Disease | Told had liver condition, past 12 months | (25) |
| Over 60 plus any risk | All people 60 and above with a health risk factor | Aggregate variable |
| Social/Demographic Risk Variables |  |  |
| Variable Name | Description | Literature |
| Can't afford care | Needed but couldn't afford medical care, past 12 months | Variable selected to support future analysis |
| No insurance | Has no health insurance coverage | Variable selected to support future analysis |
| Delayed care | Delayed medical care due to cost in last 12 months | Variable selected to support future analysis |
| Male | Selected male as gender | (21) |
| History of smoking | Has smoked more than 100 cigarettes in lifetime | (26, 28) |
| Over 60, Over 60, Over 40 | Includes all people aged 40 and above, 50 and above, and 60 and above. | (21, 29) |

We propose three stages of risk at which society can mitigate the emergence and consequences of a new disease: (1) the risk that a pathogen will spill over into the human population, (2) the risk of transmission among humans once a disease has spilled over, and (3) the risk of hospitalization or death of an infected individual. Currently, most country-wide efforts to mitigate COVID-19 focus on reducing human-to-human transmission largely through distancing policies *(9, 10)*, aimed at reducing the rate of increase in severe cases, and therefore of overwhelming hospital capacity (flattening the curve). We focus our analysis on component (3) to better understand the distribution of hospitalization and mortality risk across industries and geographical space. This may help government agencies and nonprofits target the allocation of already scarce resources (targeted control). It will also help prioritize a strategic staged geography- or occupation-based reduction of social distancing measures, and return of the workforce in the near future (national back-to-work strategy). Better planning may also increase the return on investment for public policy, keeping essential businesses functioning in high-risk industries and preventing avoidable deaths from COVID-19.

## Results

Our analysis shows that approximately 80% of the US workforce has at least one COVID-19 health risk factor, and approximately 11% of the US workforce is over 60 with at least one additional COVID-19 risk factor [Table 2]. When obesity, the most common health factor, is excluded, 56% percent of the workforce is at risk, and 10% percent of the workforce is over 60 and at risk. We also consider social and demographic variables that have the potential to interact with or may cause health risks, such as smoking history or the inability to afford medical care. For example, 11% of workers in industries deemed essential by the US government do not have health insurance (∼10 million people). A lack of health care coverage or perceived inability to afford care could prevent patients from seeking treatment early, exacerbating risk of severe outcomes *(11)*. Our results show that 8% of the total workforce, equivalent to 12 million people, delayed medical treatment in the last 12 months due to cost. As with the health variables, we err on the side of including variables with marginal significance in our analysis for added flexibility in future studies using our dataset.

**Table 2:**
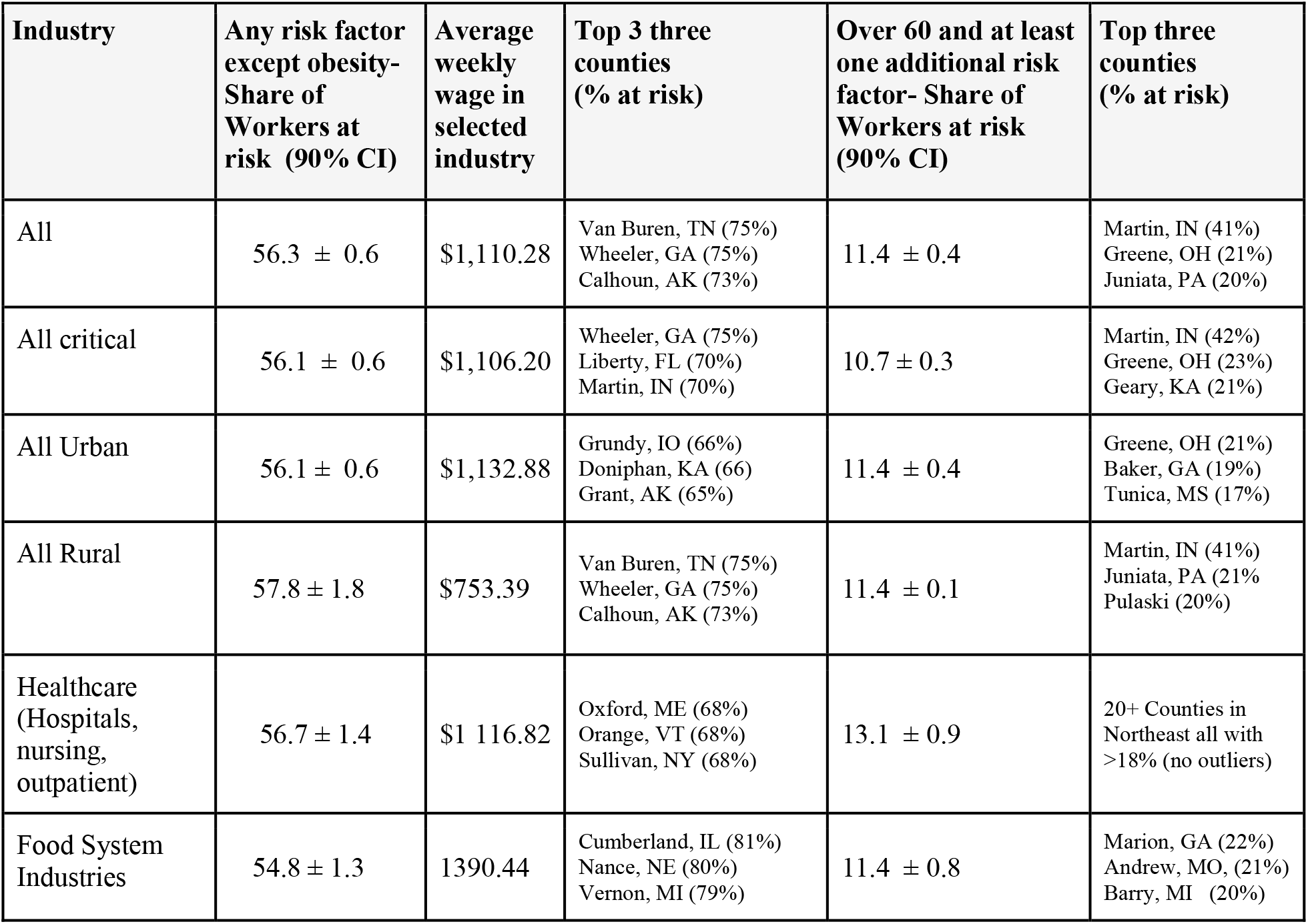
Select National Statistics

| Industry | Any risk factor except obesity- Share of Workers at risk (90% CI) | Average weekly wage in selected industry | Top 3 three counties (% at risk) | Over 60 and at least one additional risk factor- Share of Workers at risk (90% CI) | Top three counties (% at risk) |
| --- | --- | --- | --- | --- | --- |
| All | 56.3 ± 0.6 | \$1,110.28 | Van Buren, TN (75%)<br>Wheeler, GA (75%)<br>Calhoun, AK (73%) | 11.4 ± 0.4 | Martin, IN (41%)<br>Greene, OH (21%)<br>Juniata, PA (20%) |
| All critical | 56.1 ± 0.6 | \$1,106.20 | Wheeler, GA (75%)<br>Liberty, FL (70%)<br>Martin, IN (70%) | 10.7 ± 0.3 | Martin, IN (42%)<br>Greene, OH (23%)<br>Geary, KA (21%) |
| All Urban | 56.1 ± 0.6 | \$1,132.88 | Grundy, IO (66%)<br>Doniphan, KA (66)<br>Grant, AK (65%) | 11.4 ± 0.4 | Greene, OH (21%)<br>Baker, GA (19%)<br>Tunica, MS (17%) |
| All Rural | 57.8 ± 1.8 | \$753.39 | Van Buren, TN (75%)<br>Wheeler, GA (75%)<br>Calhoun, AK (73%) | 11.4 ± 0.1 | Martin, IN (41%)<br>Juniata, PA (21%)<br>Pulaski (20%) |
| Healthcare (Hospitals, nursing, outpatient) | 56.7 ± 1.4 | \$1 116.82 | Oxford, ME (68%)<br>Orange, VT (68%)<br>Sullivan, NY (68%) | 13.1 ± 0.9 | 20+ Counties in Northeast all with >18% (no outliers) |
| Food System Industries | 54.8 ± 1.3 | 1390.44 | Cumberland, IL (81%)<br>Nance, NE (80%)<br>Vernon, MI (79%) | 11.4 ± 0.8 | Marion, GA (22%)<br>Andrew, MO, (21%)<br>Barry, MI (20%) |

Spatial variation across the United States is remarkable. The percentage of the workforce with any risk factor across all industries ranges from 71% to 93%. After removing obesity from the risk factor criteria, the proportion of the population categorized as at risk declines (56%), but the spatial dispersion grows, ranging from 37% to 75%. The highest risk is concentrated in the Midwest and the lowest in the Intermountain West and West Coast. As little as 5% of the workforce over age 60 in some counties (e.g. Lander County, NV) has a risk factor, while in others, the percentage over age 60 and at-risk is over 40% (Martin County, IN). Notable disparities exist within regions, states, and between neighboring counties. For example, Greene County, Ohio’s workforce over 60 is twice as at risk than the workforce of bordering counties. This trend is found in clusters of both rural and urban counties, including those centered in Marion, IN; Pulaski, MO; and Geary, KA. Somewhat counter-intuitively, there is no significant difference in the overall proportion of the total workforce at risk between rural and urban counties. However, outlier counties with extremely high or low proportions of at-risk workers are more likely to be rural and less populated. The proportion of the workforce at risk only ranged from 46% to 66% across urban counties, while the range across rural counties was from 37% to 75%. This is perhaps due to the small number of total workers and the low diversity of industries in rural counties relative to urban regions, or disparities in income between urban and rural locales.

There is significant heterogeneity in risk between industries at the regional level, and this is even greater when broken down by county. A common trend is that sparsely populated counties are most likely to have entire industries at relatively high or low risk. This is because there are few people in those counties working in any one industry. Thus, while transmission risks may be lower in sparsely populated areas, the consequences of infection may be greater for that industry and that community. This is because critical jobs are more likely to go unfilled. In hospital and nursing industries, for example, worker absenteeism, either from increased childcare responsibilities caused by school closures or worker illness, can translate into higher patient mortality *(12)*.

The design of our data dashboard enables thousands of different combinations of geographical area, industry, and risk factor to be viewed and compared. We enable comparisons between different combinations of filters by displaying the confidence intervals for all summary statistics within the dashboard. For example, in the Northeast United States, the proportion of the healthcare workforce that is over 60 and has an additional health risk factor (16.2% ± 2.2) is on average higher than in the rest of the country (12.2% ± 0.9). In contrast, the healthcare workforce in counties in the south, mid-west, and west is more likely to delay seeking healthcare due to cost (9.3 ± .08) than in the Northeast (5.8% ±1.7). There are notable outliers in poorer counties in the west, where wages are half the national average and workers are three times as likely to delay care than in the Northeast (Fergus, MT; Fremont, CO and. This points to a need to consider the interplay of social and health risk factors in individual industries and geographical contexts when coordinating disaster response efforts or a national return-to-work policy.

Other examples of industries and areas with disproportionally high health risks compared to the surrounding areas include: the food systems industry, including transportation and warehousing services, the Texas Panhandle and South-central Florida, where the workforce is nearly twice as likely to be over 60 and at risk than the national average; crop and animal production on the Atlantic seaboard from New Jersey to Maine; and oil and gas extraction and processing in Pennsylvania. Some of these trends might not align with regional averages and instead could be the result of small workforces and idiosyncratic variation. This illustrates how the absence of a few essential workers could put entire industries or regions at risk.

## Discussion

Society failed to manage the risk of SARS-CoV-2 spilling from wildlife to people, and most countries are now struggling to reduce person-to-person spread. However, our analysis suggests countries can also mitigate overall risk by planning for and managing the consequences of infection. Developing these strategies necessitates identifying the consequences of infection at the community level. This involves understanding optimal resource allocation within hospitals and factors that influence hospital labor and materials supply *(12, 13)*. It also means planning policies to respond to the exposure of sections of the broader labor force to COVID-19 complications.

COVID-19 relief planning has focused on urban centers and areas where community transmission likely began earlier due to international travel, and is exacerbated through public transport, and tighter living quarters, such as New York City. Our analysis suggests that although rural counties in the United States might have a relatively lower rates of transmission currently, portions of their populations can have a greater risk of hospitalization and death once infected. Essential industries in rural counties are made up of smaller, and sometimes more homogenous, workforces. As such, the absence of just a small number of essential workers could lead to the collapse of these industries, leaving entire communities without necessary goods and services. For example, large meat processing plants in Iowa and Pennsylvania were shut down due to workers testing positive for the virus, and workers at a chicken processing plant in Georgia walked out in protest over being exposed to other employees with the virus *(14, 15)*. Larger industries in urban counties, on the other hand, are likely more resilient to worker absenteeism due to greater opportunity for worker substitution. As such, the small size of an industry is itself a risk factor. Consideration of this may be critical to the efficacy of policies to implement staged back-to-work policies in the US. Where rural workers are unlikely to be able to return to work, planning for enhanced mobility of replacement workers from urban centers may be beneficial. Similar strategies were adopted in China wherein essential healthcare workers were moved into Hubei to support a dearth caused by increased infection rates and a more severe lockdown *(16)*.

Social distancing measures outside of urban centers in the United States are simplified by more physically spread out communities and a lack of public transport. However, workers in essential industries, such as hospitals, food systems and public services (utilities, public safety), are less likely to be able to work from home *(17)*. Furthermore, workers in rural and urban essential industries where working from home is less frequently an option, are more likely to have social risk factors, such as lack of insurance coverage or inability to afford medical care *(11)*. This category of workers is also more likely to be in the bottom half of the income distribution, less likely to be white, and less likely to have a college degree *(11)*. Therefore, it is possible that the workers in the US most vulnerable to COVID-19 exposure are also the workers at the highest risk of hospitalization or death. The interplay of social and health risk factors will be important when considering where to focus relief efforts.

Our analysis demonstrates important heterogeneity in susceptibility to COVID-19 between geographic areas, across industries, and depending on the most pertinent health and social risk factors. A state-level analysis is likely not granular enough to efficiently allocate resources in support of a national-level policy. Currently, resources are triaged between locations based on need, a fact supported by the seizure of ventilators across New York State for use in New York City *(18)*. This is justified with COVID-19 confirmed case load being concentrated in particular localities in the United States, and as part of an effort to flatten the rate of increase of confirmed cases. However, we caution that efforts to contain the virus to urban and other hotspots may already have failed (e.g. Blaine County, Iowa), and therefore response efforts should now also consider the risk of disease severity in sectors of the population *(2)*. This requires a shift of focus to high-risk counties and industries. Failure to protect vulnerable workforces could lead to the breakdown of essential industries in these counties and cause avoidable deaths and hardship in their communities. It might also create further hotspots of viral transmission, allowing COVID-19 to resurge in cities as they manage a return-to-work strategy.

## Data Availability

all data are publicly available

## Acknowledgments

The authors contributions are as follows: Samantha Maher: data curation, formal analysis, writing- original draft preparation; Alexandra E. Hill: data curation, formal analysis, writing- review and editing, Peter Britton: data curation, formal analysis, visualization; Eli. P Fenichel: conceptualization, data curation, methodology, writing- original draft preparation; Peter Daszak: writing- review and editing; Carlos Zambrana-Torrelio: writing- review and editing; and Jude Bayham: conceptualization, methodology, data curation, formal analysis, visualization, writing- review and editing. The authors declare no competing interests. Funding for authors Maher, Daszak, and Zambrana-Torrelio from EcoHealth Alliance is provided by the The National Institute of Allergy and Infectious Diseases (NIH) and the US Agency for International Development (USAID). All data and code is available for download from the Github repository “labor_risk_dashboard” as well as directly from the dashboard itself at https://covid.yale.edu/innovation/mapping/complications/

## Supplementary Materials

Materials and Methods

Reference #31 only in Materials and Methods

---

**Fig. 1.**
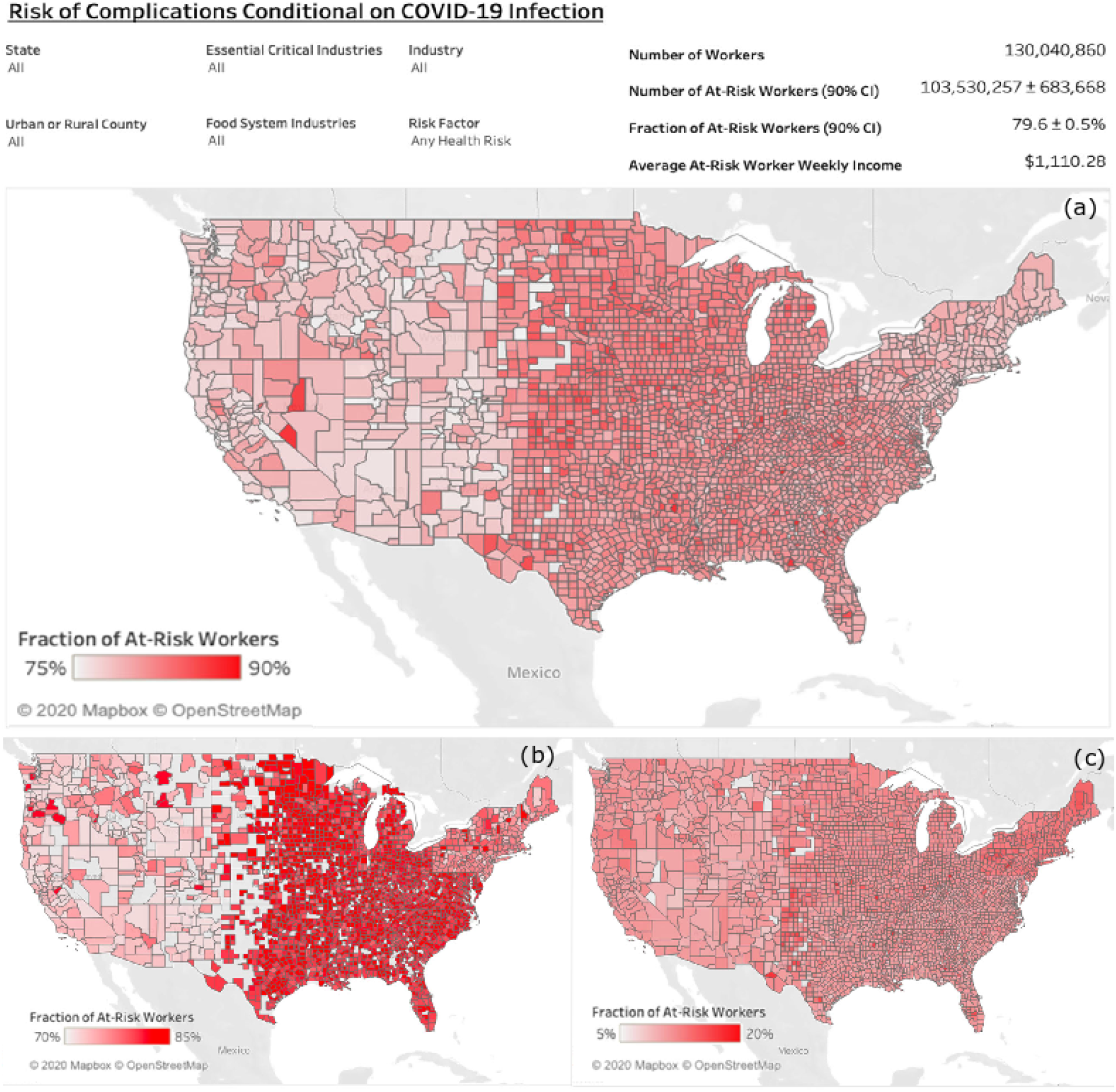
Figure 1 show results from our dashboard available at https://covid.yale.edu/innovation/mapping/complications/. (a) Proportion of at-risk workers across all industries. (b) Proportion of the healthcare workforce that has a health risk factor. (c) Proportion of the workforce across all industries that is over 60 years with an additional health risk.

## Supplementary Materials for

### Materials and Methods

We selected risk factors for complications from COVID-19 infection from the medical literature using a keyword search in MedRxiv, the preprint server for Health Sciences operated by Cold Spring Harbor Laboratory. Our keyword search consisted of the term “COVID-19” with at least one of the phrases “risk factor,” “comorbidity,” or “case fatality.” We selected relevant studies from the search results and created a list of risk factors shown to correlate with increased case severity or mortality in COVID-19 patients. We then cross-referenced the complete list with a list of health risk factors created through text- and data-mining of medical literature by the Kaggle Community, a subsidiary of Google LLC and online community of data scientists and machine learning practitioners.

We recognize that the consensus on which social, demographic, and physiological characteristics pose the greatest risk to COVID-19 patients will likely change as more data from different settings becomes available. Risk factors with marginal or inconclusive evidence (i.e. smoking, asthma, obesity) are therefore included in the list despite the small number of articles available and the dynamic nature of the data sources. The dataset includes studies made available before the end of March 2020, many of which are from the first months of the outbreak in China.

Data for individuals health and social risk factors are sourced from the National Health Interview Survey (NHIS), part of the Integrated Public Use Microdata Series, and consists of digital “information on the health, health care access, and health behaviors of the civilian, non-institutionalized U.S. population”*(6)*. We selected samples from 2016 through 2018 and matched survey variables with the list of risk factors from our literature review. With the exception of “chronic respiratory diseases” and “immunodeficiency,” all risk factors correlated with a question on the NHIS survey from 2018. Although respiratory diseases are major COVID-19 comorbidities, questions about such conditions were not explicitly asked about in the NHIS survey. We included variables for conditions leading to and encompassed by the term “chronic respiratory diseases,” including asthma, history of smoking, and lung cancer.

We pulled social and demographic variables in addition to health risk factors, including industry of employment, occupation, insurance coverage, ability to afford medical care, gender, and age. There is some evidence that male patients are significantly more likely to die from COVID-19 complications than female patients *(21)*. Similarly, patients 40 years and over show an increasing risk of mortality, with patients aged 60 and over constituting the most at-risk group *(21,29)*. In the United States, it is possible that lack of health care coverage or perceived inability to afford care will prevent patients from seeking treatment early on, leading to complications later. As with the health variables, we err on the side of including a variable in the study for added flexibility in future analysis using our dataset.

We construct a dataset in R using the microdata downloaded from the IPUMs Health Survey webpage *(6)*. We map NHIS survey responses to the presence or absence of each health and social risk factor and create additional categories for individuals of any age having at least one of the health risk factors and individuals over the age of 60 with at least one health risk factor. We use industry and occupation codes for all survey participants to sort them into working and non-working populations and create a filter for essential industries using the Department of Homeland Security’s criteria (30).

Microdata is aggregated into summary statistics for each region, industry, and occupation using the *srvyr* package in R *(24)*. First, we sum the number of survey respondents (n) in each region-industry-occupation category. We do the same for the number of people in each region-industry-occupation category for each social and health risk factor. Next, we use the sampling weight variable (sampweight) provided by IPUMs to map each survey respondent to the number of people in the United States workforce that their response represents, based on their demographic characteristics *(6)*. The region, occupation, and industry categories in IPUMS are unique and mutually exclusive for each respondent, so they can be summed to get the number of people in different permutations of region, industry, and occupation. However, each survey respondent can have multiple health and social risk factors present, and those categories are not additive. The *srvyr* package provides variance estimates based on the stratified sample design of the NHIS *(24)*.

The risk data were then merged with data on county-level employment from the Quarterly Census of Earnings and Wages (QCEW) *(7)*. We use the 2018 annual averages QCEW NAICS-based data files to obtain annual estimates of employment by industry (at the 3-digit NAICS level) for all reporting counties in the U.S. These data represent the number of workers who are covered by Unemployment Insurance for each employer in the county. This does not count self-employed workers and unpaid family workers, and might double-count workers who are employed by multiple firms within the year. Importantly, these data do not estimate the number of workers in the workforce, but rather the average number of jobs in each industry throughout the year. We use these data rather than estimates of the number of workers because they bear more significance in understanding the impacts of COVID-19 on potential disruptions to businesses and the U.S. economy, more broadly. For each county and industry, we sum employment across all ownership types (private and local, state, and federal government) to obtain one estimate of total annual employment per county and 3-digit NAICS industry. We then match each NAICS industry to the corresponding NHIS industry to combine the employment figures with data on health risk factors.

The health risk data from the NHIS is reported at the Census region while the QCEW is reported at the county. We apply the regional rates of health risk by industry to all counties within the region. However, the variance estimates are based on the population of the region, a larger geographic area than the county. We adjust the variance estimate to account for the lower precision of the regional estimate applied to the county level. Each health risk estimate is 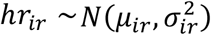. We rescale the regional statistical population (workers), *N*_*R*_, used to calculate the variance from the NHIS, by the county population of workers, *N*_*c*_:

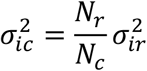

We construct 90% confidence intervals based on the county level variance estimate, 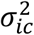, which is unambiguously larger than the region variance estimate because the county population is less than the region.

We created a dashboard and an interactive map to explore the finished data set using Tableau’s interactive data visualization software. Users are able to select geographic areas by both state and county as well as customize areas of interest at the county level. Filters are available to enable sorting and display for all combinations of industries, both essential and not essential, and additional filters are available for food system industries. All risk factors from this analysis, both social and health-related, can be selected on the dashboard. A table with summary statistics and confidence intervals updates automatically with each selection and includes the average weekly wages for all at-risk workers in each selected category. We intend for the dashboard to allow flexibility in future analysis, and anticipate its usefulness in future analyses on potential wage losses, health workforce mobility, and resource allocation.

